# Phylodynamics of Human Metapneumovirus and Evidence for a Duplication–Deletion Model in G-Gene Variant Evolution

**DOI:** 10.1101/2025.02.27.25322691

**Authors:** Stephanie Goya, Ethan B. Nunley, Preston C. Longley, Jamie R. Mathis, Christina G. Varela, Da Yae Kim, Marc Nurik, Samia N. Naccache, Alexander L. Greninger

## Abstract

**Background:** In December 2024, human metapneumovirus (hMPV) gained global attention amid rising cases in Chinese hospitals, prompting a World Health Organization (WHO) statement indicating case numbers remained within expected ranges. To assess whether a variant of public health concern was emerging and to examine global hMPV phylodynamics, we conducted a genomic surveillance study in western Washington State.

**Study design:** We sequenced hMPV genomes from inpatient and outpatient samples collected between 2021–2025 in western Washington State and constructed phylogenomic and phylodynamic trees.

**Results:** We obtained 60 hMPV-A and 39 hMPV-B genomes, including 13 from November 2024–January 2025. Following COVID-19 pandemic disruptions, hMPV seasonality returned to typical patterns after 2023. Genomic analysis showed hMPV-A predominance since 2022–23, with co-circulation of A2b1, A2b2, B1, and B2 sublineages. The A2b2 sublineage was most prevalent and all genomes carried a 111-nt G gene duplication.

Phylogenetic evidence suggests the 111-nt variant evolved from a prior 180-nt duplication via a 69-nt deletion, rather than through independent duplication events. Most sublineages showed multiple co-circulating clades, except A2b1. Phylodynamics showed recovery of global diversity after pandemic-related declines and a higher evolutionary rate in hMPV-A compared to hMPV-B. No distinct evolutionary features of heightened concern were detected.

**Conclusions:** Despite recent concerns, our findings indicate that hMPV circulation in the USA follows expected seasonal patterns, with ongoing introductions of diverse viral variants from preexisting sublineages rather than emergence of a novel variant. Continued genomic surveillance is essential, particularly as vaccine development progresses.

**Highlights:**

- hMPV seasonality in Washington State normalized by 2023 after pandemic-related disruption.
- Recent hMPV genomes show multiple introductions without evidence of a novel variant of concern.
- A2b2-111, the dominant sublineage, likely evolved from A2b2-180 variant via 69-nt G gene deletion.
- hMPV-A evolves faster than hMPV-B; global diversity declined in 2020 and rebounded post-2021.

## Background

In December 2024, human metapneumovirus (hMPV) gained international attention due to concerns over increases in cases in Chinese hospitals, prompting a World Health Organization (WHO) statement indicating case numbers were within expected ranges [1]. Discovered in 2001, hMPV is a seasonal respiratory virus spread through direct contact from fomites or aerosols, typically causing mild respiratory illness but potentially severe bronchiolitis and pneumonia, especially in infants, the elderly, and immunocompromised individuals [2]. In hematopoietic stem cell transplant patients, hMPV may lead to rapid respiratory failure and death in up to 27% of cases [3]. hMPV remains a global health threat, causing an estimated 643,000 annual hospitalizations and 7,700 deaths in children under 5 [4]. Treatment is supportive, as no licensed antivirals exist, though development of a combined mRNA vaccine for hMPV and respiratory syncytial virus (RSV) in children is underway [5,6].

hMPV is a negative-sense, single-stranded RNA virus with an approximately 13,000 nt genome encoding nine proteins, including fusion (F) and attachment (G) surface glycoproteins, which play key roles in viral entry and immune evasion. hMPV has two antigenic lineages, hMPV-A and hMPV-B, categorized into six phylogenetic sublineages: A1, A2a, A2b1, A2b2, B1, and B2 [7]. Interestingly, two independent nucleotide duplications have been reported in the same region of the G-gene in the A2b2 sublineage, resulting in 180nt (A2b2-180) and 111nt (A2b2-111) duplication variants [8,9]. However, hMPV genomic epidemiology and evolution dynamics remain extremely limited; only 21 hMPV genomes from 2024 were deposited in NCBI GenBank (Figure 1A). To identify whether hMPV is undergoing emergence of a variant of concern for public health, we conducted genomic surveillance in western Washington State (a region notable for early SARS-CoV-2 detection from China) alongside a global characterization of hMPV evolution over the last decade.

**Figure.**
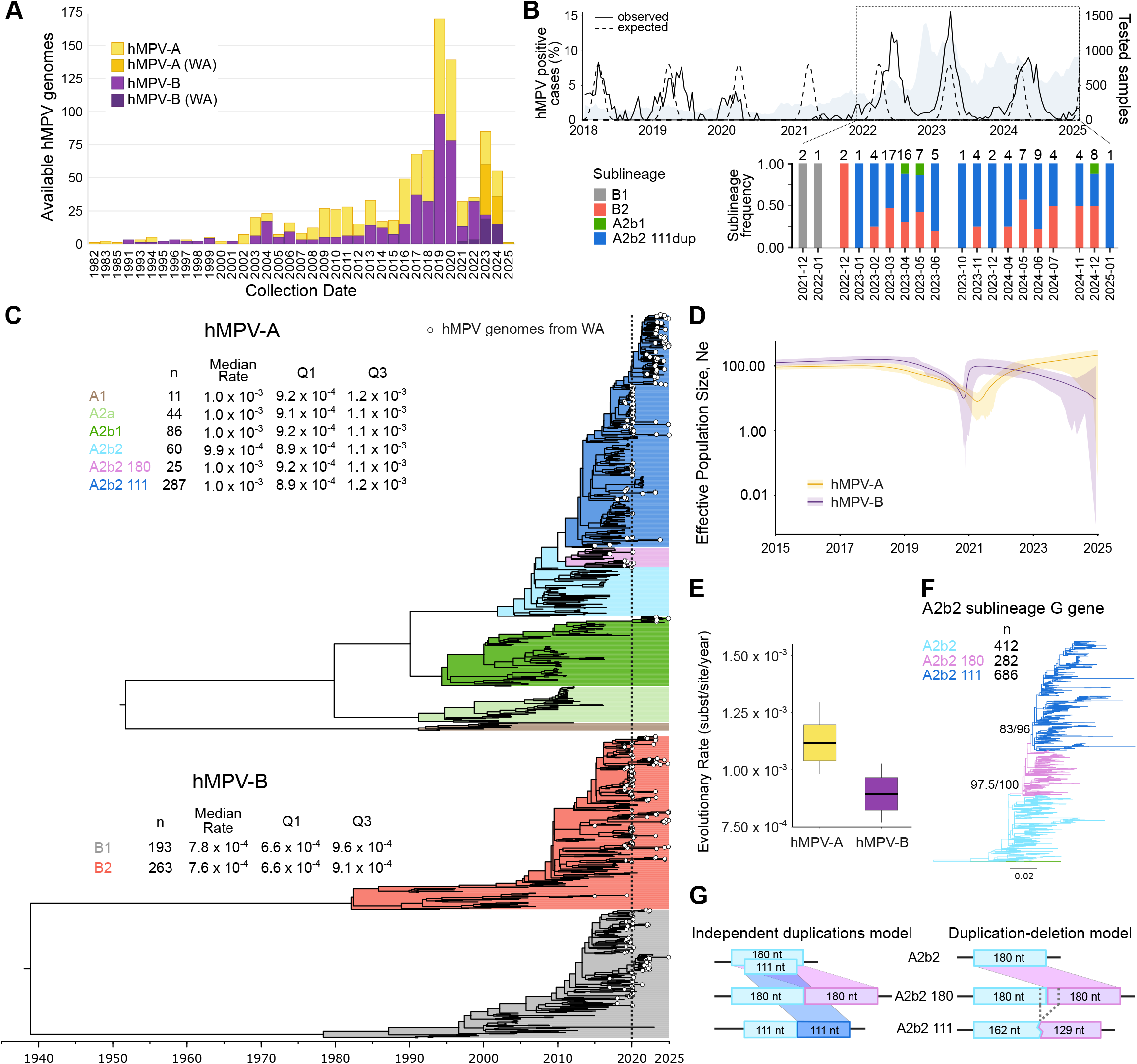
Human metapneumovirus genomic epidemiology. A) Number of hMPV genomes in NCBI GenBank by year, grouped by lineage (hMPV-A and hMPV-B). New Washington State (WA) genomes are shown in darker shades. B) Top: Biweekly hMPV positivity rates from UW Medicine (2018–2025). Solid line indicates observed positivity; dashed line represents expected seasonal pattern [17]. Background color shows total tests; the dotted box marks the study period. Bottom: Monthly frequency of WA hMPV sublineages; total genome count is shown atop each bar. C) Time-scaled Bayesian phylogenies of hMPV-A and hMPV-B. Sublineages are colored; WA genomes marked with dots. Dashed vertical line indicates 2020. Left: number of genomes, median evolutionary rate (substitutions/site/year), and interquartile range per sublineage. Scale: substitutions/site/year. D) Extended Bayesian skyline plots of hMPV-A and hMPV-B diversity pre- and post-COVID-19, based on analysis shown in panel C. E) Evolutionary rates of hMPV-A and hMPV-B. Box plots show median (horizontal line), 95% HPD interval (box), and full range (whiskers). F) Maximum-likelihood tree of A2b2 G-gene sequences. Branches are colored by sublineage; SH-aLRT/ultrafast bootstrap support shown for key nodes. Legend indicates sequence counts. G) Models of A2b2 G-gene duplication origins. Left: “Independent duplications” model proposes independent event emergence of 180-nt and 111-nt variants. Right: “Duplication–deletion” model suggests A2b2-111 arose via a 69-nt deletion from A2b2-180.

### Study design

We sequenced hMPV genomes from deidentified remnant samples collected from inpatients and outpatients in western Washington State (2021-2025). Samples tested positive from BioFire^®^ RP2.1 (LabCorp Seattle) or Hologic Panther^®^ Fusion (University of Washington Virology). BioFire^®^-tested samples were quantitated using a hMPV RT-qPCR [10]. Samples with cycle threshold (Ct) < 33 were selected to increase the likelihood of genome recovery. This study was approved by the University of Washington IRB (STUDY00000408, STUDY00000885).

RNA was extracted using the Roche MagNA^®^ Pure 96. Sequencing libraries were prepared with Illumina^®^ Respiratory Virus Enrichment Kit or QIAseq^®^ xHYB Respiratory Panel and sequenced on an Illumina NextSeq^®^2000 or NovaSeq^®^6000. Viral genomes were assembled using the REVICA-STRM pipeline (https://github.com/greninger-lab/REVICA-STRM). Consensus genomes and raw reads were deposited in NCBI under BioProject PRJNA1027483 (Supplementary Table 1).

We incorporated global hMPV genomes from NCBI GenBank (as of January 10, 2025) filtered for human-derived sequences (7-15kb length, <20% N), yielding 513 hMPV-A and 456 hMPV-B genomes. Alignments were performed with MAFFT v7.511 and phylogenies inferred using IQ-TREE v2.2 with SH-alrt and UFBoot2 support [11,12]. Phylodynamic analysis used BEAST 2.7.6 with an uncorrelated relaxed clock and Extended Bayesian Skyline model, with convergence assessed in Tracer v1.7 [13]. Maximum clade credibility trees (MCCT) were summarized with TreeAnnotator, considering clades with posterior probabilities ≥ 0.8 as supported [14].

To investigate A2b2 G-gene duplication history, we inferred expanded G gene and F gene phylogenies using an additional 945 A2b2 F gene and 1,494 G gene sequences from NCBI GenBank. Tree files and R markdown code are available at https://github.com/greninger-lab/MPV_genomics. Full methods are described in Supplementary Material.

## Results

We obtained 99 hMPV genomes (60 hMPV-A and 39 hMPV-B), including 13 recent hMPV from November 2024– January 2025 (Table 1, Figure 1A). Hospital-based surveillance revealed disrupted hMPV seasonality in Washington State in 2020, with no outbreak in 2021. A delayed outbreak peaked in mid-2022 at 11.6% test positivity, followed by a typical season in 2023 with increased positivity (14.6%) but returning to expected levels (9%) in the 2024 season (Figure 1B). Since 2022-23, hMPV-A predominated, with A2b2-111 the most frequent sublineage (Figure 1B). No significant differences in patient age or gender were observed across outbreaks or sublineages (G-test, p > 0.1).

**Table.** Demographic and epidemiological details of hMPV genomic surveillance in Washington State.

|  | hMPV-A | hMPV-B |
| --- | --- | --- |
| Total (n = 99) | 60 | 39 |
| 2021-2022 | 0 | 3 |
| 2022-2023 | 32 | 20 |
| 2023-2024 | 21 | 10 |
| 2024-2025 | 7 | 6 |
| Age Group (n = 94) |  |  |
| <5 years old | 10 | 5 |
| 5 - 18 years old | 5 | 3 |
| 19 - 65 years old | 31 | 17 |
| >65 years old | 13 | 10 |
| Sex (n= 94) |  |  |
| Female | 38 | 18 |
| Male | 21 | 17 |
| hMPV median Ct value [IQR] | 25.30 [7.05] | 24.12 [4.67] |
| Clinical Status (n = 69) |  |  |
| Outpatient | 32 | 17 |
| Inpatient | 12 | 8 |

Phylodynamic analysis showed genetically diverse recent hMPV strains are associated with multiple introductions into Washington State (Figures 1C). An exception was A2b1, where post-2020 strains clustered monophyletically. Global hMPV diversity (measured as effective population size) declined during 2020, likely reflecting pandemic-related transmission bottlenecks, with recovery after 2021 in hMPV-B and then hMPV-A (Figure 1D). Estimated evolutionary rate was higher for hMPV-A (1.11×10^−3^ substitutions/site/year, 95% High Posterior Density: 1.03×10^−3^ - 1.19×10^−3^) than hMPV-B (8.93×10^−4^ substitutions/site/year, 95% HPD: 7.69×10^−4^ - 1.02×10^−3^) (Figure 1E). However, no rate differences were observed among sublineages (Figure 1C) or pre-/post-2020 clades (Supplementary Material).

Time-scaled phylogenies suggest hMPV-A and hMPV-B have circulated for approximately 74 and 86 years, respectively (Figure 1C). The most prevalent sublineage currently, A2b2, has circulated since 2002. However, all post-2020 A2b2 strains carried a 111-nt G-gene duplication. To provide more resolution to the A2b2 history, expanded G-gene phylogeny revealed that A2b2-180 variant emerged first, and then the A2b2-111 variant diverged from the A2b2-180 clade (Figure 1F, Supplementary Figure 1). This pattern remained consistent after trimming the duplication copy from the G-gene alignment to control for artifacts (Supplementary Figure 2). Analysis of the alignment suggested A2b2-111 may have arisen via a 69-nt deletion from the A2b2-180 variant, rather than through independent duplication events (Figure 1G). Analysis of hMPV sequences derived from only complete genomes or expanded F-gene phylogeny was consistent with either successive duplication-deletion or independent duplication events (Figure 1C, Supplementary Figure 3). Nucleotide similarity also indicates that A2b2-111 is more similar to A2b2-180 strains (96.4%, IQR: 95.6-97%) than that original A2b2 strains (94.7%, IQR: 93.6-95.5%) (Supplementary Material).

## Discussion

Despite increasing global awareness of hMPV outbreaks, genomic epidemiology remains very limited with 82% of available genomes from the last five years originating from the USA. Containment measures during the COVID-19 pandemic disrupted hMPV transmission globally, causing reduced circulation, diminished viral diversity, and altered seasonality. In Washington State, seasonality returned to pre-pandemic patterns (season from January to June) by 2023-24, aligning with national surveillance data, including a delayed 2022 outbreak and increased positivity rate during 2023 [15].

Phylodynamic analysis showed that hMPV-A is evolving faster than hMPV-B. Although hMPV has been circulating for over 70 years, all current hMPV genomes belong to four sublineages (A2b1, A2b2, B1, B2). Post-2020 most sublineages showed multiple co-circulating clades, except A2b1. A2b2 was dominant in our study, with all recent strains carrying a 111-nt G gene duplication [7,16]. While prior studies described the 111-nt and 180-nt duplications as independent evolutionary events, our complete G-gene phylogeny suggests a duplication-deletion model, in which the A2b2-111 variant arose from a 69-nt deletion of the A2b2-180 variant. Nucleotide similarity and tree structure support this model and may reflect selective pressures balancing immune evasion and structural fitness.

In summary, pandemic-related containment measures significantly affected hMPV transmission and diversity, though the virus has since recovered historical seasonality. Although currently circulating strains of hMPV do not exhibit any distinctive genomic features that warrant heightened concern, seasonal circulation and continuous evolution should be monitored closely, especially as vaccine trials are underway. Limitations in our study include a sample set biased toward high viral loads and lack of clinical outcome data. Expanded surveillance, including clinical metadata and broader geographic sampling is needed to better understand hMPV evolution and its implications for public health.

## Data Availability

hMPV consensus genomes are available at NCBI GenBank under the accession numbers PV052133-PV052231. Sequencing reads are available associated with NCBI BioProject PRJNA1027483. Supplementary Table 1 details the hMPV specimen metadata and including accession numbers to NCBI.

https://github.com/greninger-lab/MPV_genomics

## Acknowledgments

ALG reports contract testing from Abbott, Cepheid, Novavax, Pfizer, Janssen, and Hologic, outside of the described work. All the other authors declare no conflict of interest. This research did not receive any specific grant from funding agencies in the public, commercial, or not-for-profit sectors.

## CRediT authorship contribution statement

Conceptualization (SG, ALG), Methodology & Resources (EBT, PCL, JRM, CGV, DYK, MK), Data curation & Formal analysis (SG, EBT, PCL, JRM), Funding acquisition (SNN, ALG), Validation (SNN, ALG), Writing – original draft (SG, ALG), Writing – review and editing (all authors).

