## Supplementary Figure 2 for "Phylodynamics of Human Metapneumovirus and Evidence for a Duplication–Deletion Model in G-Gene Variant Evolution"

**Supplementary Figure 2. Maximum-likelihood phylogenetic tree of the hMPV A2b2 sublineage G gene (with duplication copy removed).** The tree includes 1,191 non-duplicate complete or partial sequences (≥400 nt) from GenBank (as of Jan 10, 2025), with three A2b1 sequences as outgroup. To assess bias, the gene region corresponding to the duplication copies was removed from to alignment. Branches are colored by sublineage. Sequences with unexpected G gene structures (e.g., absent or incorrect duplications) are marked with colored circles at the tips. Tip labels show GenBank accession, country, and collection date, and "CG" indicates sequences from complete genomes. SH-aLRT/Ultrafast bootstrap values are shown for key nodes. The scale bar represents nucleotide substitutions per site.

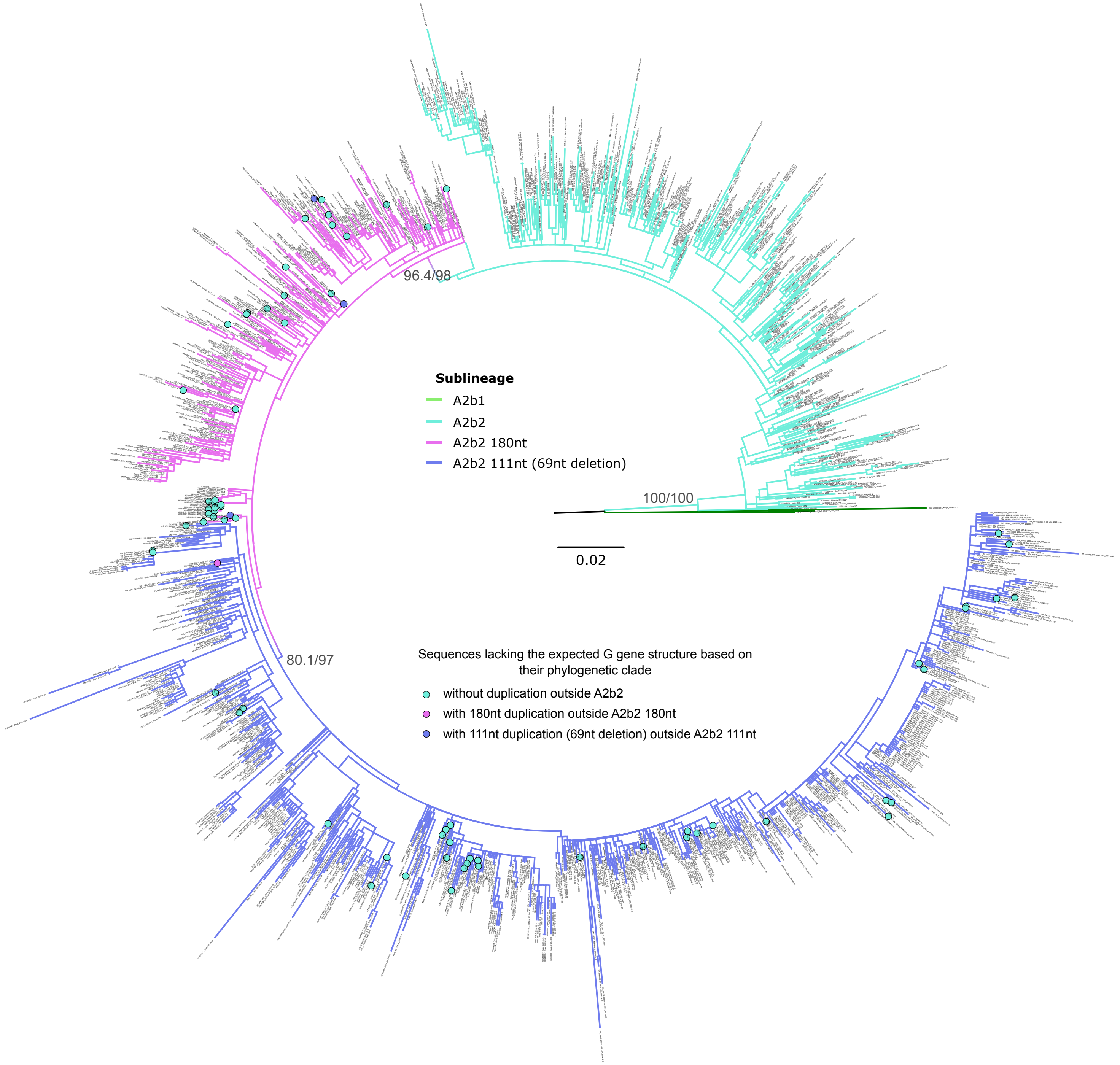
