## Supplementary Figure 3 for "Phylodynamics of Human Metapneumovirus and Evidence for a Duplication–Deletion Model in G-Gene Variant Evolution"

**Supplementary Figure 3. Maximum-likelihood phylogenetic tree of the hMPV A2b2 sublineage F gene.** The tree includes 749 non-duplicate complete or partial sequences (≥400 nt) from GenBank (as of January 10, 2025), with three A2b1 sequences included as an outgroup. Branches are colored by sublineage. Tip labels show the GenBank accession number, country, and sample collection date; “CG” indicates sequences from complete genomes and those lacking the G gene duplication but clustering within duplication-associated clades are marked with light blue circles at the tips. Sequences downloaded as F gene only, therefore unable to confirm the presence or absence of G gene duplication, are marked with red circles. SH-aLRT and Ultrafast bootstrap support values are shown for key nodes. The scale bar represents nucleotide substitutions per site.

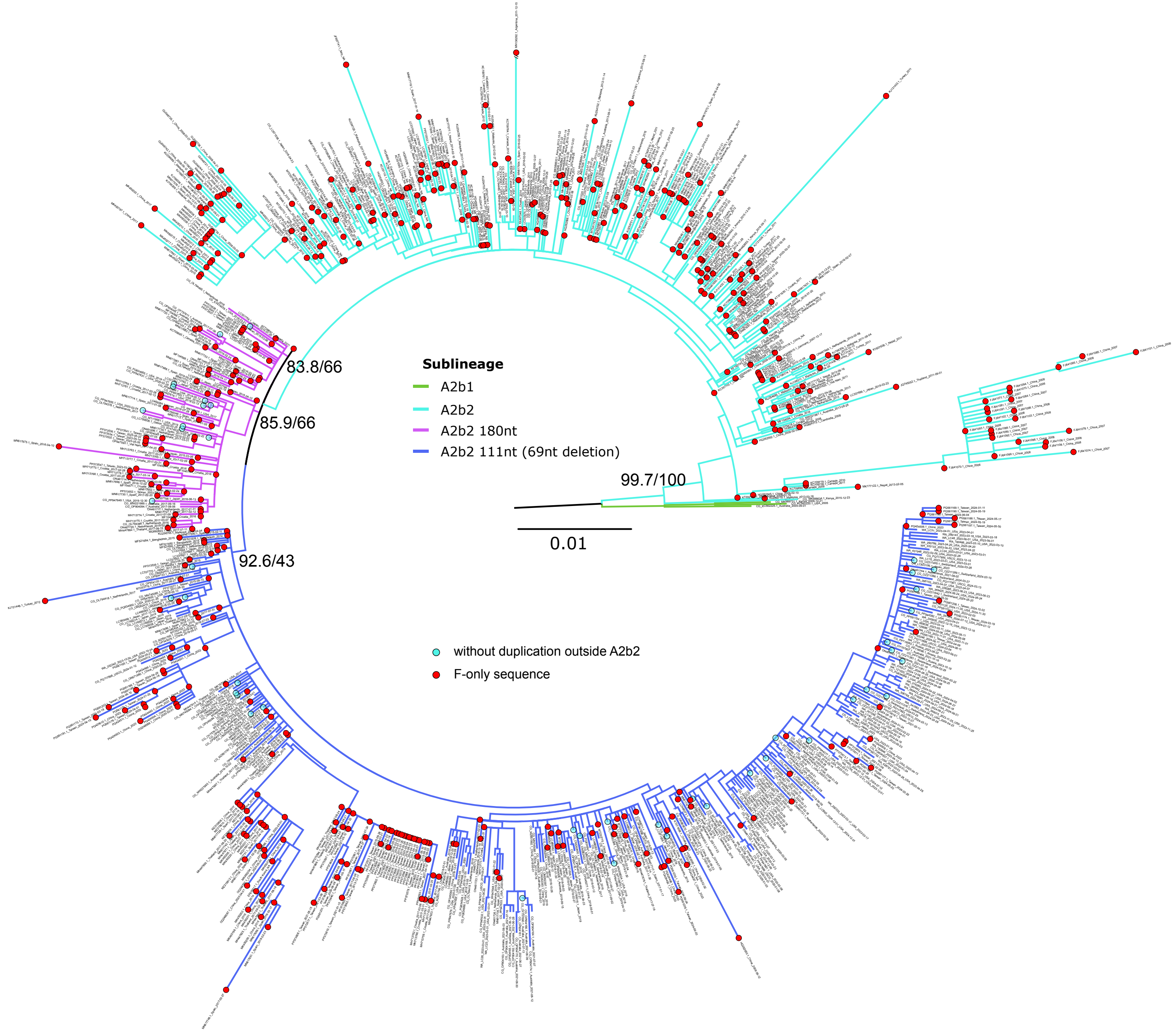
